# The Acute Effects of Lipoprotein Apheresis on Monocyte Subset Distribution and Transcriptome in Patients With and Without Elevated Lipoprotein(a)

**DOI:** 10.1101/2025.06.08.25328552

**Authors:** Brian T. Dinh, Isidoro Cobo, Julie-Ann Dutton, Mark McClellan, Jacob Rendler, Lizhu Lin, Mia Murphy, Patrick M. Moriarty, Calvin Yeang

**Author notes:** Corresponding author: Brian Dinh.

## Abstract

Lipoprotein apheresis effectively lowers low density lipoprotein and lipoprotein(a) extracorporeally. Both low density lipoprotein and lipoprotein(a) are causal risk factors for cardiovascular disease and interact with monocytes in the circulation, although the latter is the predominant lipoprotein carrier of pro-inflammatory oxidized phospholipids and considered to be a much more potent direct activator of monocytes. While an anti-inflammatory effect on monocytes may be expected with lipoprotein apheresis treatment based on its effect of acutely lowering atherogenic lipoproteins, it is plausible that monocytes may also be activated as they transit through the extracorporeal system. In this study, the net effect of lipoprotein apheresis acutely on monocytes collected from 18 patients with elevated lipoprotein(a) [median pre-treatment level of 449.0 nmol/L] and 10 patients with elevated low density lipoprotein-cholesterol (and without elevated lipoprotein(a) [median pre-treatment level of 4.1 nmol/L]) was evaluated using immunophenotyping and transcriptomic analyses. Lipoprotein apheresis acutely reduced Lp(a) and LDL-C by >70%. In both groups, lipoprotein apheresis shifted the monocyte subset distribution towards an expansion of CD14^+^ CD16^-^ classical monocytes. The number of significantly up- and down-regulated genes in monocytes from patients with elevated lipoprotein(a) was more than 3-fold more than monocytes from those without elevated lipoprotein(a). Both pro- and anti-inflammatory gene expression pathways were represented following a lipoprotein apheresis session regardless of pre-treatment lipoprotein(a) levels.

## Introduction

Atherosclerotic cardiovascular disease (ASCVD) is driven by vascular inflammation initiated by excess levels of apolipoprotein B (apoB)-containing lipoproteins, including low density lipoprotein (LDL) and lipoprotein(a) [Lp(a)]^1–3^. Accumulation of apoB-containing lipoproteins, and their subsequent oxidation within the arterial wall promotes endothelial injury and activation, resulting in activation and recruitment of circulating monocytes and tissue-resident macrophages into the developing atherosclerotic plaque^4^. There, they can further differentiate into foam cells and play critical roles in the propagation of chronic inflammation within the arterial wall^5,6^.

In contrast to LDL, which undergoes oxidative modification largely *in situ*, Lp(a) uniquely carries pro-inflammatory oxidized phospholipids that are covalently bound to its apo(a) component and embedded its lipid phase, making it an intrinsically pro-inflammatory lipoprotein in the circulation^7–10^. Consistent with its role as the major lipoprotein carrier of oxidized phospholipids, targeted and potent Lp(a) lowering by an investigational therapy reduced monocyte pro-inflammatory gene expression, in contrast to potent LDL lowering with PCSK9 monoclonal antibody treatment^11,12^.

While there are currently no clinically available pharmacologic therapies that potently lower Lp(a), lipoprotein apheresis (LA) is an FDA approved treatment for patients with high risk for recurrent ASCVD events due to elevated Lp(a) and/or familial hypercholesterolemia. During each LA treatment session, the patient’s plasma is passed through an adsorption column that removes apoB-containing lipoproteins, resulting in an acute 70-80% reduction in LDL-cholesterol (LDL-C) and Lp(a)^13^. Observational studies suggest reduced ASCVD event rates on LA treatment, however the lack of randomized, sham controlled outcomes studies and few mechanistic studies on LA limit a complete understanding of this therapy^14,15^. In this study, we evaluated the effect of LA on the monocyte subset distribution and transcriptome in patients with ASCVD who have elevated and non-elevated Lp(a) levels.

## Methods

### Study Population

Patients treated with bi-weekly dextran sulfate absorption LA with the LA-15 system (Kaneka Corp., Minato City, Tokyo, Japan) for clinical indications at the University of Kansas Medical Center were enrolled between 2021-2022. Blood samples from each patient were collected immediately before and after one LA treatment session. Plasma measurements of LDL-C and Lp(a) with the Tina-quant assay (Roche Diagnostics, Indianapolis, IA, USA) were performed under standard of clinical care by the clinical laboratory at the University of Kansas Medical Center and as a send out test to the Mayo Clinic, respectively. Peripheral blood mononuclear cells (PBMCs) were sent to the University of California San Diego for analyses. This study was approved by both the University of Kansas Medical Center and University of California San Diego Human Research Protections Programs.

### PBMC Collection

PBMCs collection was performed as described elsewhere^16^. Briefly, vacutainer cell preparation tubes with sodium heparin (CPT) (BD Biosciences, San Jose, CA, USA) were used to collect 8 mL of whole blood from patients immediately pre- and post-LA, gently inverted, and centrifuged at 1,800g for 20 minutes at room temperature (RT). The mononuclear cell layer was collected and resuspended to a 50 mL volume of RPMI medium (Corning, Corning, NY, USA). Following centrifugation at 300g for 10 minutes, the supernatant was discarded and the cell pellet was resuspended with 15 mL of RPMI + 12.5% human serum albumin (HSA) (Gemini Bio, Sacramento, CA, USA) + 1 μM flavopiridol (Millipore Sigma, Burlington, MA, USA). Pre-LA samples were left at RT for 2-3 hours prior to being processed with post-LA samples simultaneously. 15 mL of RPMI + 11.25% HSA + 1 μM flavopiridol + 10% DMSO (Invitrogen, Carlsbad, CA, USA) was added dropwise to each tube and gently inverted. Resuspended PBMCs were then placed in a pre-chilled Mr. Frosty (Thermo Scientific, Waltham, MA, USA) for 12 hours at −80°C prior to storage in a cryobox at −80°C.

### Sample preparation, Flow activated cell sorting (FACS), and flow cytometry analysis

Frozen PBMCs pre- and post-LA were thawed at 37°C, diluted with an equal volume of PBS + 2 mM EDTA, and passed through a 70 µM cell strainer (Biopioneer, San Diego, CA, USA). Following centrifugation at 400g for 7 minutes, the supernatant was discarded, and the cell pellet was resuspended in PBS + 2 mM EDTA. Zombie NIR (Biolegend, San Diego, CA, USA) and RNasin (Promega, Madison, WI, USA) were added as per manufacturers’ instructions and left to incubate for 20 minutes in the dark. Samples were then incubated with Human Trustain FcX (Biolegend, San Diego, CA, USA) for 10 minutes at RT, followed by staining with FITC-CD3, FITC-CD19, FITC-CD56, FITC-66b, APC-HLA-DR, PerCP/Cyanine5.5 CD14, and BV421-CD16 (Biolegend, San Diego, CA, USA) for 30 minutes at 4°C in the dark as per manufacturer’s instructions. Following 2 washes with PBS + 2 mM EDTA + 2.5% Ultrapure BSA (Molecular Cloning Lab, San Francisco, CA, USA), cells were passed through a 70 µM cell strainer and resuspended in 200 µL PBS + 2mM EDTA. FACS was performed using a FACSAris II (BD Biosciences, San Jose, CA, USA) by the Flow Cytometry Core at the San Diego Center for Aids Research and monocytes were sorted directly into Trizol LS (Invitrogen, Carlsbad, CA, USA). The gating strategy is illustrated in the supplemental files (**Supplemental Figure 1).** Monocyte subsets were evaluated with FlowJo v10.9 (BD Biosciences, San Jose, CA, USA).

### Library Preparation and RNA-Sequencing

Library preparation and RNA-sequencing was performed as described elsewhere^17–20^. Briefly, sorted monocytes were lysed in Trizol LS followed by RNA isolation using the Direct-zol RNA MicroPrep kit (Zymo Research, Tustin, CA, USA). 500 ng – 1 µg total RNA was used for poly-A tailed RNA enrichment with Oligo d(T) Magnetic Beads (New England Biolabs, Ipswich, MA, USA) followed by fragmentation in 2X Superscript III first-strand buffer containing 10 mM DTT (Invitrogen, Waltham, MA, CA) for 9 minutes at 94°C. Subsequently, 10 µL fragmented mRNA, 0.5 µL Random Primers (Invitrogen, Waltham, MA, CA), 0.5 µL Oligo dT primer (Invitrogen, Waltham, MA, CA), 0.5 µL SUPERase-In (Ambion, Austin, TX, USA), 1 µL of 10 mM dNTPs and 1 µL of 10 mM DTT were incubated at 50°C for three minutes. After incubation, 5.8 µL nuclease-free water, 1 µL of 100 mM DTT solution, 0.1 µL of 2 µg/µL Actinomycin D (Sigma-Aldrich, St. Louis, MO, USA), 0.2 µL of 1% Tween-20 (Sigma-Aldrich, St. Louis, MO, USA), and 0.2 µL of Superscript III (Invitrogen, Waltham, MA, CA) were incubated in a PCR machine under the following protocol: 25°C for 10 minutes, 50°C for 50 minutes, followed by a 4°C hold. The resulting product was then purified using RNAClean XP beads (Beckman Coulter, Brea, CA, USA) according to the manufacturer’s instructions. The RNA/cDNA double-stranded hybrid was then added to 1.5 µL of Blue Buffer (Enzymatics, Beverly, MA, USA); 1.1 µL of dUTP mix consisting of 10 mM dATP, dCTP, dGTP, and 20 mM dUTP; 0.2 µL of RNAse H (5 U/µL); 1 µL of 10 U/ µL DNA polymerase I (Enzymatics, Beverly, MA, USA); 0.15 µL of 1% Tween-20; and 1.05 µL of nuclease-free water. Following incubation at 16°C for 2.5 hours, the resulting dsDNA was purified using 28 µL of Speed Bead Magnetic Carboxylate (GE Healthcare, Chicago, IL, USA) diluted with 20% PEG 8000:2.5M NaCl to final concentration of 13% PEG. This was followed by washing twice with 80% ethanol, air drying, and elution in 40 μL of 0.05% Tween-20. The resulting purified dsDNA underwent end-repair by blunting followed by A-tailing and adapter ligation as described elsewhere using Bioo Barcodes (Bioo Scientific, Austin, TX, USA), IDT TruSeq Unique Dual Indexes or Kapa Unique Dual-Indexed Adapters, 15 μL Rapid Ligation Buffer (Enzymatics, Beverly, MA, USA), 0.33 μL 1% Tween-20, and 0.5 μL T4 DNA ligase HC (Enzymatics, Beverly, MA, USA) [39]. Libraries were PCR amplified for 11–15 cycles using Solexa IGA (AATGATACGGCGACCACCGA) and Solexa IGB primers (CAAGCAGAAGACGGCATACGA), purified using 1 μL of Speed Bead Magnetic Carboxylate in 15.2 μL of 20% PEG 8000:2.5 M NaCl, washed with 80% ethanol, and eluted in 0.05% Tween-20. Eluted libraries were quantified using a Qubit dsDNA HS Assay Kit and sequenced on an Illumina NovaSeq 6000 (Illumina, San Diego, CA, USA). Sequencing quality metrics are included as a supplementary table (**Table S1**).

### RNA-Sequencing Analysis

Monocyte transcriptome data analysis was performed as previously described^17,18^. Briefly, reads were mapped to the hg38 reference genome using STAR with default parameters. Transcript quantification was performed with HOMER analyzeRepeats.pl under the following parameters: - condenseGenes, -count exons, and -noadj. Expression values for each transcript were calculated using the Homer analyzeRepeats.pl with the following parameters: -condenseGenes, -count exons, and -tpm. Differential expression analysis was calculated using the HOMER getDiffExpression.pl tool with the following parameter: FDR <0.05. Only libraries with >75% mapped reads and >7 million uniquely mapped reads were used for downstream analysis. A full list of differentially expressed genes (DEGs) is provided in the supplemental files (**Tables S2-S5)**. Significant DEGs with P <0.05 and a Log2FC <-0.25 or >0.25 were uploaded to Metascape for Canonical and Hallmark pathway enrichment analyses^21^.

### Data Analysis

All statistical analyses were performed using SPSS v29.0.2 (SPSS Inc., Chicago, IL, USA). Monocyte subset data is represented as mean percentage (IQR). Comparison of monocyte subset distribution between elevated and non-elevated Lp(a) groups used independent two-sided T-Tests. Comparison of monocyte subsets between pre-LA and post-LA within the elevated or non-elevated Lp(a) groups utilized paired T-Tests. Statistics comparing patient characteristics utilized independent T-Tests for numerical variables and Chi-squared tests of independence for categorical variables. Box and whisker plots were generated using R v4.3.0 using the ggplot2 package^22^. Volcano plots were generated using R v4.3.0 using the following packages: tidyverse, rcolorbrewer, and ggrepel^22–25^.Venn diagrams were made with DeepVenn [46]. Pathway enrichment analyses were performed using Metascape^26^.

## Results

### Patient Characteristics

Thirty patients undergoing LA treatment on a biweekly basis, 3 of whom were LA naïve (**Table 1**). Two patients were excluded from the final analysis due to poor RNA-sequencing QC parameters and therefore 18 and 10 patients were included in the elevated and non-elevated Lp(a) groups respectively. Within the elevated Lp(a) group, median (IQR) Lp(a) and LDL-C levels immediately pre-LA treatment were 449.0 (300.1-570.3) nmol/L and 78.0 (65.0-118.5) mg/dL, respectively. In this group, LA yielded a median 77.4% reduction in Lp(a) and a 73.7% reduction in LDL-C. In the non-elevated Lp(a) group, pre-LA Lp(a) and LDL-C levels were 5.7 (0.6-21.3) nmol/L and 197.0 (164.8-216.0) mg/dL, respectively. LA resulted in an acute lowering of LDL-C by 74.4% while Lp(a) were not quantified post-LA due to the limited assay accuracy in this expected very low Lp(a) range.

**Table 1:** Study cohort characteristics and medical history.

|  | **Elevated Lp(a)** | **Non-Elevated Lp(a)** | **P-Value** |
| --- | --- | --- | --- |
| N | 18 | 10 |  |
| Age, mean (SD) | 61 (7.7) | 70 (7.0) | P < 0.01 |
| Female, n (%) | 10 (56) | 8 (80) | P > 0.05 |
| LA-Naïve, n (%) | 2 (11) | 1 (10) | P > 0.05 |
| Pre-LA Lp(a) nmol/L, median (IQR) | 449.0 (300.1-570.3) | 4.1 (<7.0-70.0)* | P < 0.00001 |
| Post-LA Lp(a) nmol/L, median (IQR) | 89.0 (56.6-113.1) |  |  |
| % Lp(a) reduction, mean (SD) | 77.4 (9.0) |  |  |
| Pre-LA LDL-C mg/dL, median (IQR) | 78.0 (65.0-118.5) | 198.0 (164.8-216.0) | P < 0.001 |
| Post-LA LDL-C mg/dL, median (IQR) | 23.0 (15.0-25.0) | 50.5 (38.3-58.8) | P < 0.001 |
| % LDL-C reduction, mean (SD) | 73.7 (8.3) | 74.4 (7.0) | P > 0.05 |
| **Medications** |  |  |  |
| Statins | 10 | 1 | P < 0.05 |
| PCSK9i | 7 | 1 | P > 0.05 |
| Niacin | 1 | 0 | P > 0.05 |
| Ezetimibe | 7 | 1 | P > 0.05 |
| Omega-3 FA | 6 | 5 | P > 0.05 |
| Antiplatelets | 15 | 8 | P > 0.05 |
| **Medical History** |  |  |  |
| Diabetes | 9 | 3 | P > 0.05 |
| History of MI | 5 | 1 | P > 0.05 |
| Revascularization | 12 | 3 | P > 0.05 |
| Stroke | 2 | 0 | P > 0.05 |
| Carotid Disease | 1 | 3 | P > 0.05 |
| Stent | 12 | 2 | P < 0.05 |
| CABG | 5 | 2 | P > 0.05 |
| TIA/stroke | 3 | 2 | P > 0.05 |

\*Due to the lower limit of quantification of Lp(a) of 7 nmol/L, the pre-LA Lp(a) levels in the non-elevated Lp(a) group are expressed as median (range). Post-LA Lp(a) levels were below the limit of quantification in this group.

### Monocyte Subset Distribution in Patients Undergoing LA

Classical (CM), intermediate (IM), and non-classical (NCM) monocyte subsets were evaluated by flow cytometry in patients with elevated and non-elevated Lp(a) levels before and after LA (**Table 2**). No significant difference in the monocyte subset distribution was observed between the elevated vs non-elevated Lp(a) groups pre-LA (P = 0.65, 0.45, and 0.98 for the CM, IM, and NCM subsets respectively; **Figure 1A**). However, both the elevated and non-elevated Lp(a) groups demonstrated a significant shift favoring CM post-LA alongside a proportional decrease in the IM and NCM subsets (P < 0.05 for all subsets; **Figure 1B/C)**.

**Figure 1:**
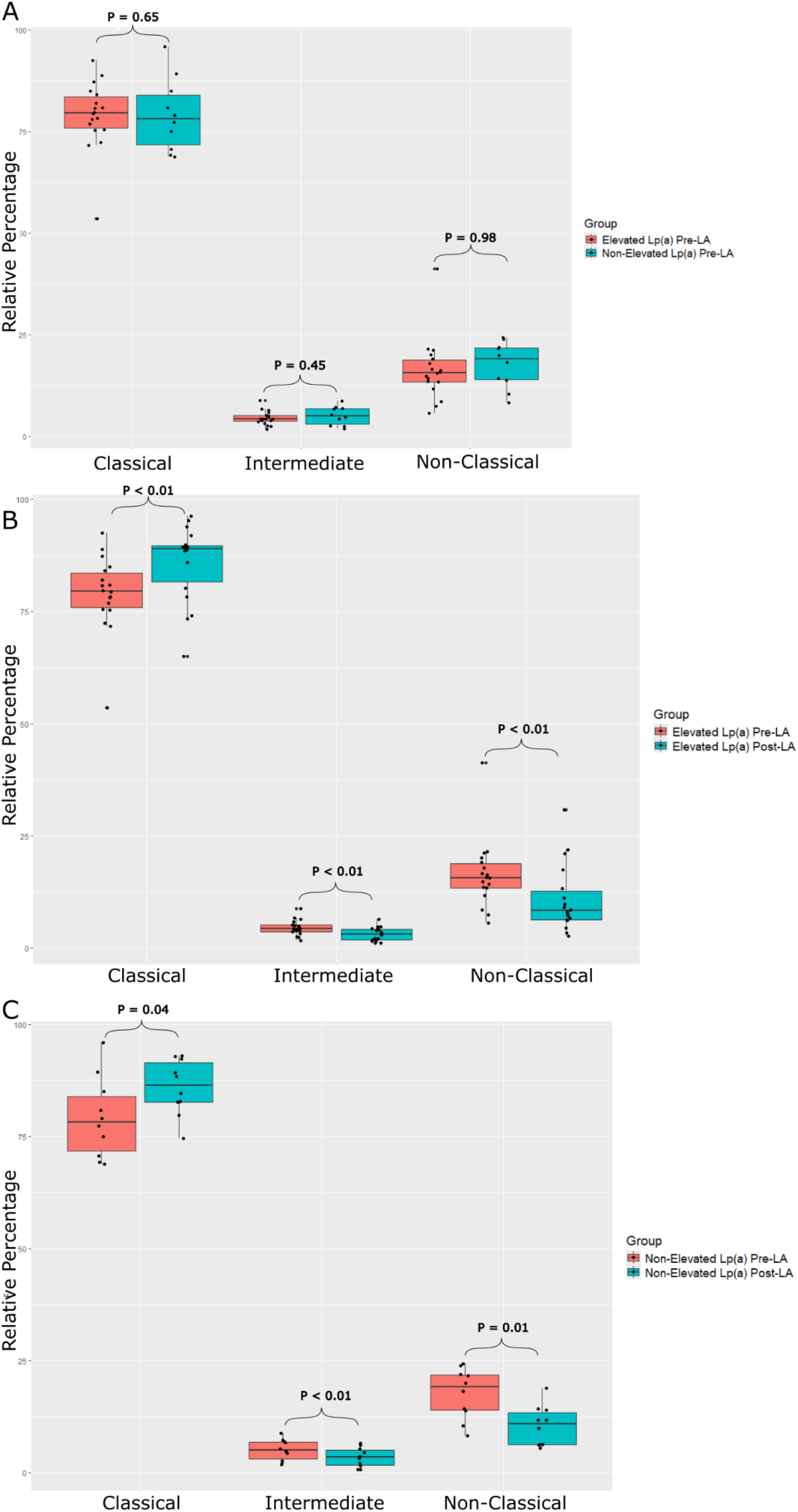
Box and whisker plots comparing the relative monocyte subset distribution in (**A**) elevated and non-elevated Lp(a) patients pre-LA, (**B**) elevated Lp(a) patients pre-LA and post-LA, and (**C**), non-elevated Lp(a) patients pre-LA and post-LA.

**Table 2:** Relative monocyte subset distribution in elevated and non-elevated Lp(a) patients pre- and post-LA treatment. The data are presented as mean (interquartile range) or mean (SD)

|  | Elevated Lp(a) (%) | P-Value | Non-Elevated Lp(a) (%) | P-Value |
| --- | --- | --- | --- | --- |
| **Pre-LA Classical (CD14+ CD16-)** | 79.0 (75.5 – 84.3) | $P < 0.01$ | 79.1 (70.4 – 86.1) | $P < 0.01$ |
| **Post-LA Classical (CD14+ CD16-)** | 85.9 (80.0-90.3) |  | 86.0 (82.0 – 92.4) |  |
| **% Change Classical (Post-LA vs Pre-LA)** | 6.9 (4.6) |  | 6.9 (5.7) |  |
| **Pre-LA Intermediate (CD14+ CD16+)** | 4.6 (3.5 – 5.4) | $P < 0.01$ | 5.1 (2.6 – 7.0) | $P < 0.01$ |
| **Post-LA Intermediate (CD14+ CD16+)** | 3.1 (1.8 – 4.3) |  | 3.5 (1.4 – 5.5) |  |
| **% Change Intermediate (Post-LA vs Pre-LA)** | -1.4 (1.1) |  | -1.6 (0.8) |  |
| **Pre-LA Non-Classical (CD14+ CD16-)** | 16.4 (13.0 – 19.4) | $P < 0.01$ | 17.7 (13.0 – 22.4) | $P < 0.01$ |
| **Post-LA Non-Classical (CD14+ CD16-)** | 10.9 (6.3 – 14.4) |  | 10.5 (6.3 – 14.1) |  |
| **% Change Non-Classical (Post-LA vs Pre-LA)** | -5.5 (4.2) |  | -7.2 (5.0) |  |

### Bulk RNA-Sequencing of Monocytes in Patients Undergoing LA

Next, bulk RNA-sequencing was performed to characterize the effect of LA on the pan-monocyte transcriptome. No significant DEGs were found comparing the elevated Lp(a) and non-elevated Lp(a) cohorts pre-LA (**Figure S2**). LA treatment was associated with 1,251 upregulated and 1400 downregulated DEGs in patients with elevated Lp(a) (**Figure 2A**) compared to 361 upregulated and 330 downregulated DEGs in those with non-elevated Lp(a) (**Figure 2B**). A full list of DEGs is listed in the supplemental files (**Tables S2-S9)**. Notably, the majority of the non-elevated Lp(a) DEGs are shared with the elevated Lp(a) group (**Figure 3**). Several genes associated with immune activation including *CLEC5A*, *TNFSF14*, *CCR2*, *CXCR1*, and *IRAK2,* were identified amongst the top 20 upregulated significant DEGs in the elevated Lp(a) group post-LA (**Table S6)**. In line with the elevated Lp(a) results, *CLEC5A* is identified in the top 20 upregulated significant DEGs alongside *TLR2* and *TLR6* in the top 30 DEGs in the non-elevated Lp(a) group (**Table S8**).

**Figure 2:**
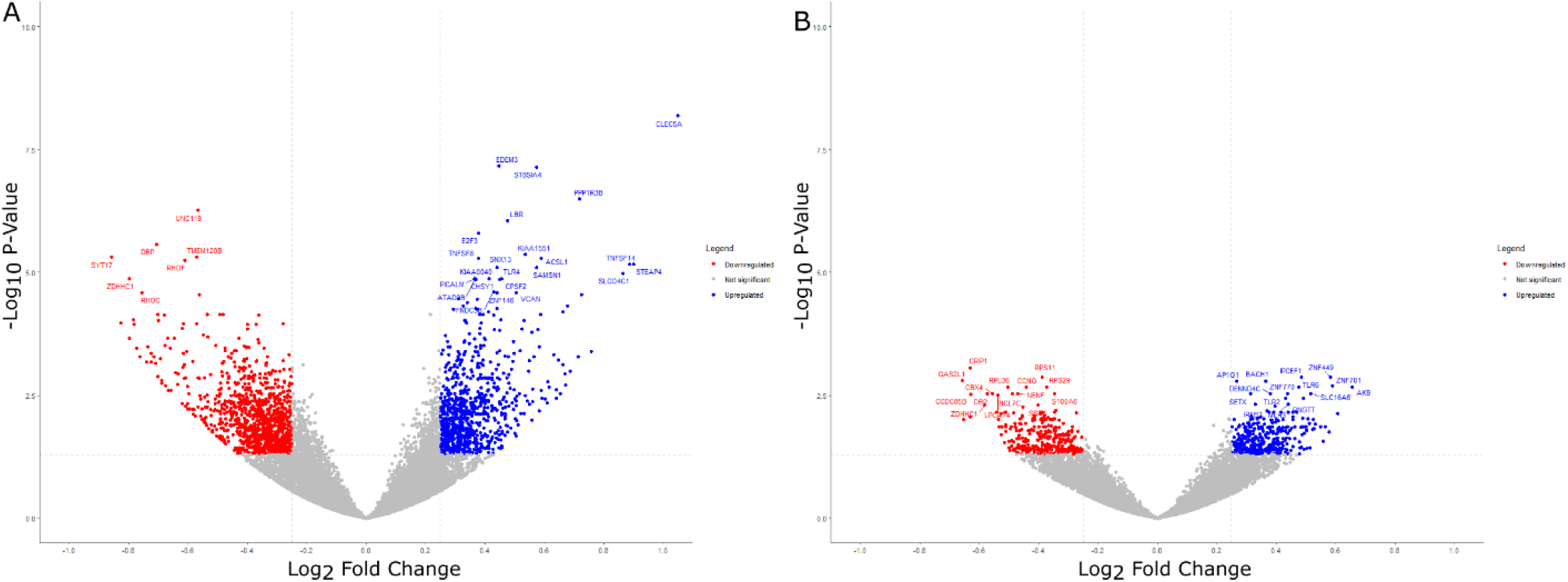
Volcano plots displaying significant DEGs in monocytes isolated from elevated (**A**) and non-elevated Lp(a) patients post-vs pre-LA (**B**).

**Figure 3:**
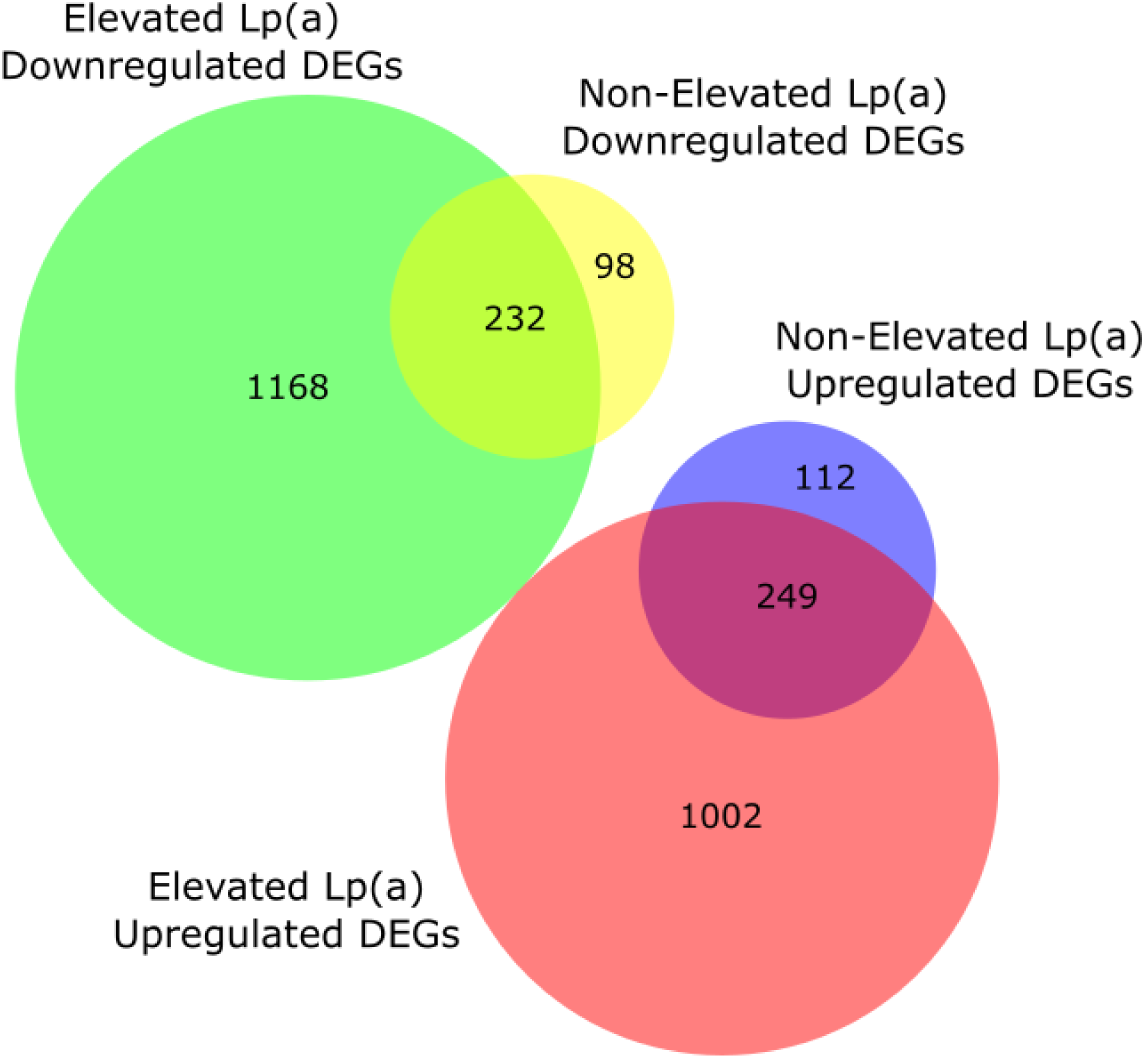
Venn diagram displaying the shared number of upregulated and downregulated DEGs between elevated and non-elevated Lp(a) monocytes pre- and post-LA. Created with DeepVenn^26^.

Hallmark & Canonical pathways enrichment analyses were performed on the upregulated and downregulated DEGs in both elevated and non-elevated Lp(a) groups (**Figure 4**)^21^. Pathways related to immune activation and inflammation were upregulated in both the elevated (**Figure 4A**) and non-elevated (**Figure 4C**) Lp(a) groups. Pathways relating to oxidative phosphorylation and DNA repair were downregulated in both the elevated (**Figure 4B**) and non-elevated (**Figure 4D**) Lp(a) group. The Hallmark Interferon Gamma Response, Apoptosis, and Fatty Acid Metabolism pathways were present in both the elevated Lp(a) upregulated DEGs and the non-elevated Lp(a) downregulated DEGs (**Figure 4E**).

**Figure 4:**
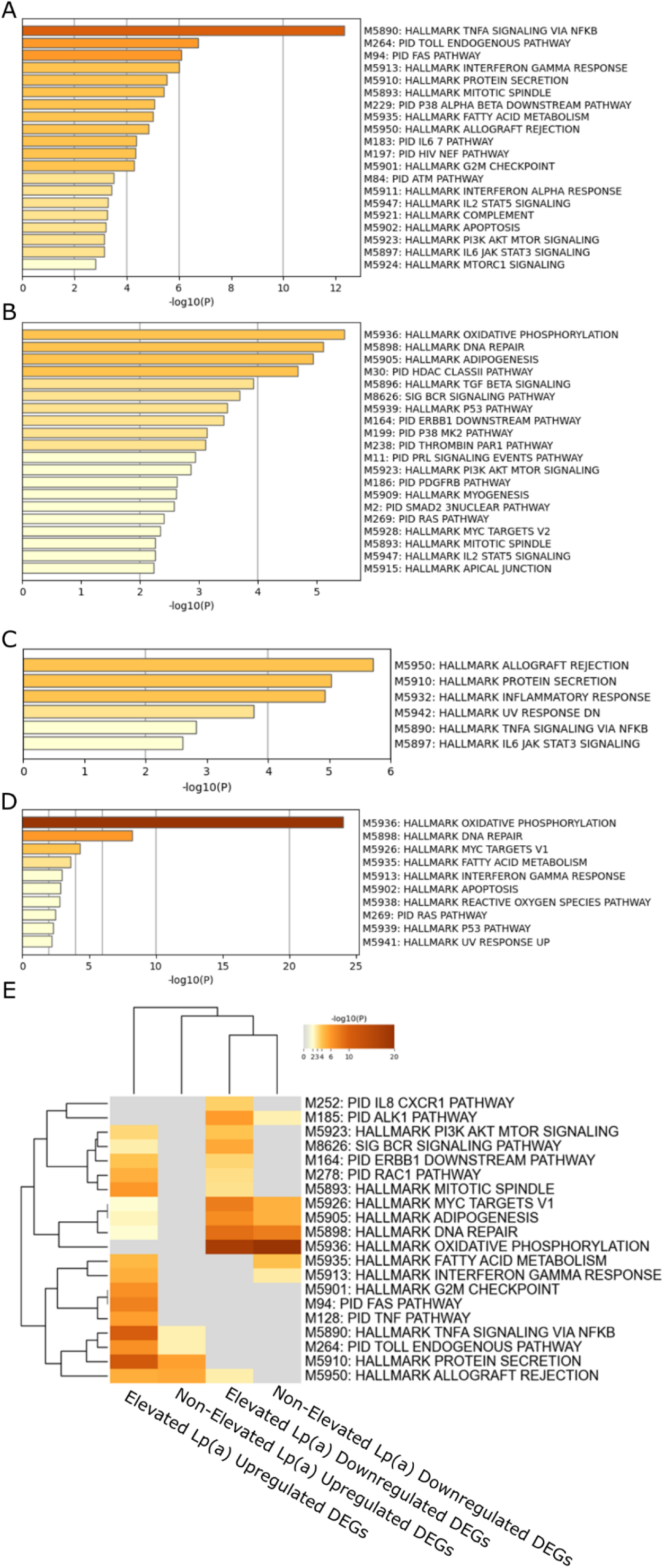
Hallmark & Canonical pathway enrichment analysis for upregulated DEGs in elevated Lp(a) patients (**A**), downregulated DEGs in elevated Lp(a) patients (**B**), upregulated DEGs in non-elevated Lp(a) patients (**C**), downregulated DEGS in non-elevated Lp(a) patients (**D**), alongside an overview comparing shared pathways (**E**).

## Discussion

Our study provides several novel observations on acute effects of LA on monocyte subsets and transcriptomic profiles in ASCVD patients with and without elevated Lp(a) levels. First, LA shifts the monocyte distribution towards classical monocytes and away from intermediate and non-classical monocytes. Second, the monocyte transcriptomic response to LA is more robust patients with elevated Lp(a), with 3-4-fold more significantly up- or down-regulated genes compared to patients with non-elevated Lp(a). Third, pathway analysis of differentially expressed genes following LA revealed both shared and unique pathways between patients with and without elevated Lp(a).

Changes in the distribution of monocyte subsets on LA, using the LA-15 system, has implications for cardiovascular disease risk. Human monocytes can be divided into different subsets based on relative CD14 vs CD16 surface expression levels: classical (CD14^+^ CD16^-^), intermediate (CD14^+^ CD16^+^), and non-classical (CD14^-^ CD16^+^)^28–30^. Classical monocytes are the most abundant in circulation and are primarily involved in the initiation of inflammatory responses. Non-classical monocytes have been shown to patrol the vascular endothelium, are involved in tissue repair, and have elevated antigen presentation gene expression. Intermediate monocytes exhibit features of both classical and non-classical subsets but are considered pro-inflammatory due to their production of reactive oxygen species and inflammatory cytokines.

In this study, LA treatment increased the relative proportion of classical monocytes and decreased the proportion of the non-classical and intermediate subsets in patients with and without elevated Lp(a), suggesting a less inflammatory monocyte phenotype. However, associations between each of the monocyte subsets and ASCVD events remain controversial and require further investigation in well-powered studies^31–41^.

Prior work by Jellinghaus and colleagues found that LA treatment resulted in an increase in the CM and a decrease in the NCM subsets, with the IM subset unchanged^42^. Another study by Walther et al. found a relative decrease in both the NCM and IM subsets with no change in the CM subset in LA-naïve patients^43^. Notably, these studies utilized patients treated with a diverse array of LA modalities, including the HELP, LF, Monet, Therasorb, Liposorber D, and DALI systems. These systems use different methods of lipoprotein removal, including the use of whole blood perfusion, which may result in significant differences in monocyte phenotype^44^.

To our knowledge, this study is one of the largest evaluating the effect of LA on monocyte subsets and the first to describe the effect of LA on monocyte gene expression. There was a general upregulation of inflammatory gene expression pathways immediately following an LA treatment session. However, monocytes from patients with elevated Lp(a) experienced a larger number of significant differentially expressed genes compared to those with non-elevated Lp(a). Additionally, upregulation of several pathways that were downregulated in the those with non-elevated Lp(a) was observed in those with elevated Lp(a), including the Hallmark Interferon Gamma Response, Apoptosis, and Fatty Acid Metabolism pathways. These findings highlight the unique roles that Lp(a) and LDL have on monocytes and suggest that monocytes isolated from elevated Lp(a) patients post-LA are more strongly activated by LA than those from non-elevated Lp(a) patients.

However, previous work by Stiekema and colleagues reported a decrease in pro-inflammatory gene expression in monocytes collected from patients treated with potent apo(a)-targeted antisense therapies^11^. These findings align with the known pro-inflammatory properties of Lp(a) and with prior genetic studies predicting that potent Lp(a) lowering would reduce CVD risk^45,46^. One explanation for the discrepancy between this study and others is that LA treatment may result in shear stress and damage, altering the monocyte phenotype, and leading to the activation of pro-inflammatory pathways. Prior work found that different LA methods altered monocyte subsets different ways, likely owing to differences in filter materials^42^. It remains to be verified whether LA is sufficient to induce inflammatory gene expression in monocytes across different modalities. Monocytes from patients with elevated Lp(a) levels are known to be trained and have increased ability to transmigrate and secrete proinflammatory cytokines^47^. While differences before stimulation may be minimal, trained monocytes from elevated Lp(a) patients may more strongly upregulate inflammatory pathways when stimulated by LA. An alternative explanation is that LA treatment is known to remove than just lipoproteins from plasma leading to pleiotropic effects^48–51^. The LA-15 system utilizes a negatively charged dextran sulfate column, which binds positively charged proteins including those on lipoproteins. It’s plausible that the removal of unidentified plasma components contributes to the increase in pro-inflammatory gene expression.

The novel observations from this study will require validation by functional monocyte assessments. An understanding of the temporal changes in monocyte subset distributions and gene expression in between bi-weekly LA sessions will be important to fully contextualize the net effects of LA. Lastly, the findings regarding the effects of LA on monocytes need to be considered in the context of the known long term cardiovascular benefits of apoB-containing lipoprotein lowering and observational studies supporting improved cardiovascular outcomes with LA therapy.

## Limitations

The analyses of monocytes immediately pre- and post-LA informs the acute effects of LA. However, samples in between bi-weekly treatment sessions were not available for analysis to inform the full spectrum of temporal changes in monocytes with chronic and repeated LA treatment. It is plausible that the observed pro-inflammatory effect of LA on monocytes is transient, and a different gene expression profile becomes dominant as the mechanical effects of LA wane while LDL and Lp(a) levels remain lower than pre-LA levels in between LA sessions.

Because the absolute number of monocytes was not available, the observed changes in monocyte subset distribution and transcription can be explained by changes in the circulating pool of monocytes without a change in the number of monocytes, or secondary to an influx of bone marrow derived or demarginalized monocytes. Finally, the magnitude of gene expression change was relatively limited, reflecting only modest differences. Future studies utilizing larger sample sizes and ex-vivo assays are necessary to validate these findings.

## Data Availability

All data produced in the present study are available upon reasonable request to the authors

## Acknowledgements

We would like to express gratitude to the patients who volunteered for this study. This publication includes data generated at the UC San Diego IGM Genomics Center utilizing an Illumina NovaSeq 6000 that was purchased with funding from a National Institutes of Health SIG grant (#S10 OD026929).

## References

1. Borén, J. et al. Low-density lipoproteins cause atherosclerotic cardiovascular disease: pathophysiological, genetic, and therapeutic insights: a consensus statement from the European Atherosclerosis Society Consensus Panel. Eur. Heart J. 41, 2313–2330 (2020).

2. Björkegren, J. L. M. & Lusis, A. J. Atherosclerosis: Recent developments. Cell 185, 1630–1645 (2022).

3. Zhang, X., Sessa, W. C. & Fernández-Hernando, C. Endothelial Transcytosis of Lipoproteins in Atherosclerosis. Front. Cardiovasc. Med. 5, (2018).

4. Gimbrone, M.A.; García-Cardeña, G. Endothelial Cell Dysfunction and the Pathobiology of Atherosclerosis. Circ Res 2016, 118, 620–636, doi:10.1161/CIRCRESAHA.115.306301. - Google Search. https://www.google.com/search?q=Gimbrone%2C+M.A.%3B+Garc%C3%ADa-Carde%C3%B1a%2C+G.+Endothelial+Cell+Dysfunction+and+the+Pathobiology+of+Atherosclerosis.+Circ+Res+2016%2C+118%2C+620%E2%80%93636%2C+doi%3A10.1161%2FCIRCRESAHA.115.306301.&rlz=1C1VDKB_enUS1005US1005&oq=Gimbrone%2C+M.A.%3B+Garc%C3%ADa-Carde%C3%B1a%2C+G.+Endothelial+Cell+Dysfunction+and+the+Pathobiology+of+Atherosclerosis.+Circ+Res+2016%2C+118%2C+620%E2%80%93636%2C+doi%3A10.1161%2FCIRCRESAHA.115.306301.&gs_lcrp=EgZjaHJvbWUyBggAEEUYOdIBBzExM2owajeoAgCwAgA&sourceid=chrome&ie=UTF-8.

5. Gui, Y., Zheng, H. & Cao, R. Y. Foam Cells in Atherosclerosis: Novel Insights Into Its Origins, Consequences, and Molecular Mechanisms. Front. Cardiovasc. Med. 9, (2022).

6. Poznyak, A. V., Nikiforov, N. G., Starodubova, A. V., Popkova, T. V. & Orekhov, A. N. Macrophages and Foam Cells: Brief Overview of Their Role, Linkage, and Targeting Potential in Atherosclerosis. Biomedicines 9, 1221 (2021).

7. Stewart, C. R. et al. CD36 ligands promote sterile inflammation through assembly of a Toll-like receptor 4 and 6 heterodimer. Nat. Immunol. 11, 155–161 (2010).

8. Kadl, A. et al. Oxidized phospholipid-induced inflammation is mediated by Toll-like receptor 2. Free Radic. Biol. Med. 51, 1903–1909 (2011).

9. Chávez-Sánchez, L. et al. Activation of TLR2 and TLR4 by minimally modified low-density lipoprotein in human macrophages and monocytes triggers the inflammatory response. Hum. Immunol. 71, 737–744 (2010).

10. Schnitzler, J. G. et al. Short-term regulation of hematopoiesis by lipoprotein(a) results in the production of pro-inflammatory monocytes. Int. J. Cardiol. 315, 81–85 (2020).

11. Stiekema, L. C. A. et al. Potent lipoprotein(a) lowering following apolipoprotein(a) antisense treatment reduces the pro-inflammatory activation of circulating monocytes in patients with elevated lipoprotein(a). Eur. Heart J. 41, 2262–2271 (2020).

12. Bergmark, C. et al. A novel function of lipoprotein [a] as a preferential carrier of oxidized phospholipids in human plasma. J. Lipid Res. 49, 2230–2239 (2008).

13. Parker, T. S. Dextran-sulfate cellulose adsorption for lowering Lp[a]. Chem. Phys. Lipids 67–68, 331–338 (1994).

14. Schettler, V. J. J. et al. The German Lipoprotein Apheresis Registry-Summary of the eleventh annual report. Atherosclerosis 398, 118601 (2024).

15. Leebmann, J. et al. Lipoprotein Apheresis in Patients With Maximally Tolerated Lipid-Lowering Therapy, Lipoprotein(a)-Hyperlipoproteinemia, and Progressive Cardiovascular Disease. Circulation 128, 2567–2576 (2013).

16. Dinh, B. et al. Isolation and Cryopreservation of Highly Viable Human Peripheral Blood Mononuclear Cells From Whole Blood: A Guide for Beginners. J. Vis. Exp. JoVE e66794 (2024) doi:10.3791/66794.

17. Cobo, I. et al. DNA methyltransferase 3 alpha and TET methylcytosine dioxygenase 2 restrain mitochondrial DNA-mediated interferon signaling in macrophages. Immunity 55, 1386–1401.e10 (2022).

18. Cobo, I. et al. Monosodium urate crystals regulate a unique JNK-dependent macrophage metabolic and inflammatory response. Cell Rep. 38, (2022).

19. Gosselin, D. et al. An environment-dependent transcriptional network specifies human microglia identity. Science 356, eaal3222 (2017).

20. Seidman, J. S. et al. Niche-Specific Reprogramming of Epigenetic Landscapes Drives Myeloid Cell Diversity in Nonalcoholic Steatohepatitis. Immunity 52, 1057–1074.e7 (2020).

21. Metascape provides a biologist-oriented resource for the analysis of systems-level datasets | Nature Communications. https://www.nature.com/articles/s41467-019-09234-6.

22. Wickham, H. Ggplot2: Elegant Graphics for Data Analysis. (Springer-Verlag New York, 2016).

23. R Core Team R: A Language and Environment for Statistical Computing. R Foundation for Statistical Computing.

24. Slowikowski, K. et al. Ggrepel: Automatically Position Non-Overlapping Text Labels with “Ggplot2”. (2024).

25. Neuwirth, E. ColorBrewer Palettes 2022. (2022).

26. Hulsen. DeepVenn -- a Web Application for the Creation of Area-Proportional Venn Diagrams Using the Deep Learning Framework Tensorflow.Js. (2022).

27. Wickham, H. et al. Welcome to the Tidyverse. J. Open Source Softw. 4, 1686 (2019).

28. Kapellos, T. S. et al. Human Monocyte Subsets and Phenotypes in Major Chronic Inflammatory Diseases. Front. Immunol. 10, (2019).

29. Boyette, L. B. et al. Phenotype, function, and differentiation potential of human monocyte subsets. PLOS ONE 12, e0176460 (2017).

30. Williams, H. et al. Monocyte Differentiation and Heterogeneity: Inter-Subset and Interindividual Differences. Int. J. Mol. Sci. 24, 8757 (2023).

31. Ruder, A. V., Wetzels, S. M. W., Temmerman, L., Biessen, E. A. L. & Goossens, P. Monocyte heterogeneity in cardiovascular disease. Cardiovasc. Res. 119, 2033–2045 (2023).

32. Kim, K.-W., Ivanov, S. & Williams, J. W. Monocyte Recruitment, Specification, and Function in Atherosclerosis. Cells 10, 15 (2021).

33. Yang, J., Zhang, L., Yu, C., Yang, X.-F. & Wang, H. Monocyte and macrophage differentiation: circulation inflammatory monocyte as biomarker for inflammatory diseases. Biomark. Res. 2, 1 (2014).

34. Hamers, A. A. J. et al. Human Monocyte Heterogeneity as Revealed by High Dimensional Mass Cytometry. Arterioscler. Thromb. Vasc. Biol. 39, 25–36 (2019).

35. Berg, K. E. et al. Elevated CD14++CD16-monocytes predict cardiovascular events. Circ. Cardiovasc. Genet. 5, 122–131 (2012).

36. Monocyte-Chemoattractant Protein-1 Levels in Human Atherosclerotic Lesions Associate With Plaque Vulnerability | Arteriosclerosis, Thrombosis, and Vascular Biology. https://www.ahajournals.org/doi/10.1161/ATVBAHA.121.316091?doi=10.1161/ATVBAHA.121.316091.

37. Rogacev, K. S. et al. CD14++CD16+ Monocytes Independently Predict Cardiovascular Events: A Cohort Study of 951 Patients Referred for Elective Coronary Angiography. J. Am. Coll. Cardiol. 60, 1512–1520 (2012).

38. Cappellari, R. et al. Shift of monocyte subsets along their continuum predicts cardiovascular outcomes. Atherosclerosis 266, 95–102 (2017).

39. Ozaki, Y. et al. Association of Toll-Like Receptor 4 on Human Monocyte Subsets and Vulnerability Characteristics of Coronary Plaque as Assessed by 64-Slice Multidetector Computed Tomography. Circ. J. 81, 837–845 (2017).

40. Afanasieva, O. I. et al. The Association of Lipoprotein(a) and Circulating Monocyte Subsets with Severe Coronary Atherosclerosis. J. Cardiovasc. Dev. Dis. 8, 63 (2021).

41. Zawada, A. M. et al. Monocyte heterogeneity in human cardiovascular disease. Immunobiology 217, 1273–1284 (2012).

42. Jellinghaus, S. et al. Lipoprotein apheresis influences monocyte subpopulations. Atheroscler. Suppl. 30, 108–114 (2017).

43. Repeated lipoprotein apheresis and immune response: Effects on different immune cell populations - Walther - 2022 - Therapeutic Apheresis and Dialysis - Wiley Online Library. https://onlinelibrary.wiley.com/doi/10.1111/1744-9987.13886.

44. Therapeutic Apheresis for Management of Lp(a) Hyperlipoproteinemia | Current Atherosclerosis Reports. https://link.springer.com/article/10.1007/s11883-020-00886-0.

45. Burgess, S. et al. Association of LPA Variants With Risk of Coronary Disease and the Implications for Lipoprotein(a)-Lowering Therapies: A Mendelian Randomization Analysis. JAMA Cardiol. 3, 619–627 (2018).

46. Dzobo, K. E., Kraaijenhof, J. M., Stroes, E. S. G., Nurmohamed, N. S. & Kroon, J. Lipoprotein(a): An underestimated inflammatory mastermind. Atherosclerosis 349, 101–109 (2022).

47. Oxidized Phospholipids on Lipoprotein(a) Elicit Arterial Wall Inflammation and an Inflammatory Monocyte Response in Humans | Circulation. https://www.ahajournals.org/doi/full/10.1161/CIRCULATIONAHA.116.020838.

48. Leebmann, J. et al. Lipoprotein Apheresis in Patients With Maximally Tolerated Lipid-Lowering Therapy, Lipoprotein(a)-Hyperlipoproteinemia, and Progressive Cardiovascular Disease. Circulation 128, 2567–2576 (2013).

49. Schettler, V. J. J. & Schettler, E. Beyond cholesterol—pleiotropic effects of lipoprotein apheresis. (2022).

50. Marlęga-Linert, J. et al. Lipoprotein apheresis affects the concentration of extracellular vesicles in patients with elevated lipoprotein (a). Sci. Rep. 14, 2762 (2024).

51. Sinzinger, H., Steiner, S. & Derfler, K. Pleiotropic effects of regular lipoprotein-apheresis. Atheroscler. Suppl. 30, 122–127 (2017).

